# Repositioned chloroquine and hydroxychloroquine as antiviral prophylaxis for COVID-19: A protocol for rapid systematic review of randomized controlled trials

**DOI:** 10.1101/2020.04.18.20071167

**Authors:** Raymond Chang, Wei-Zen Sun

**Author notes:** Corresponding author: Raymond Chang.

## Abstract

Since the SARS-CoV-2 outbreak rapidly evolved into a pandemic, there is an urgent need for rapid development, identification and confirmation of efficacious antiviral prophylaxis. In this setting, the existing drugs chloroquine (CQ) and hydroxychloroquine (HCQ) which has suggestive evidence of efficacy against SARS-CoV-2 infection and COVID-19 disease has become prime candidates to be repositioned as therapeutic and preventative agents, and a growing number of clinical trials have been registered to study their preventative potential for at-risk populations using a range of dosing schemes and outcome measures. This rapid systematic review protocol aims to provide streamlined and timely synthesis on methodologies and results of randomized controlled trials assessing the efficacy of CQ and HCQ in hopes that this will constructively inform further research as well as public health policy.

## BACKGROUND

Since its reported outbreak in late 2019, SARS-CoV-2 virus causing corona virus 2019 (COVID-19) disease has exploded from a few people suffering a respiratory disease to a pandemic of over a million cases. Current methods of infection control is largely confined to public and personal health measures, while vaccine development maybe as much as 18 months away from deployment (Higgins-Dunn, 2020).

Besides vaccination, antiviral prophylaxis is the other major pharmaceutical intervention that can be effective. Antivirals are potentially prophylactic and has been successfully applied pre- and post-exposure against viral infections (De Clercq, 2013). In the case of COVID-19, the repositioning the old anti-malarial chloroquine (CQ) and its derivative hydroxychloroquine (HCQ) has received much attention as they have *in-vitro* efficacy against SARS-CoV-2 (Wang et al., 2020) and have some preliminary evidence of clinical efficacy against COVID-19 (Gao et al., 2020; Gautret et al., 2020). The two agents have thus been proposed as potential prophylaxis against SARS-CoV-2 and COVID-19 (Chang & Sun, 2020; Nicola & Esposito, 2020) but they are nevertheless untested in the prophylaxis setting, and multiple trials targeting different populations with a total proposed enrolment of over a hundred thousand subjects world-wide using a range of doses as well as outcomes have already been registered to date.

Notwithstanding a living systematic review protocol has been proposed to assess clinical trials for COVID-19 (Maguire & Guérin, 2020), and published systematic reviews evaluating CQ and HCQ’s role as treatment for COVID-19 (Kapoor & Kapoor, 2020) or assessing clinical trials using CQ and HCQ as treatment while specifically excluding prevention studies (Rana & Dulal, 2020), there is as yet no protocol for systematically assessing clinical trials that address the preventative efficacy of CQ and HCQ against SARS-CoV-2 and COVID-19 and we aim to fulfil the gap.

As a choice of review methodology, we decided on a rapid systematic review because a traditional systematic review with its concomitant rigours of methodology can take up to two years to conduct which may be inordinately long to be informative to researchers and policy makers in the face of a rapidly evolving pandemic. Whereas rapid reviews would be a streamlined form of knowledge synthesis geared to be informative in a timely manner (Khangura et al., 2012), and would be more suitable to the subject and context of this review.

## METHODS

### Protocol and registration

This manuscript complies with the ‘Preferred Reporting Items for Systematic reviews and Meta-Analyses’ (PRISMA) guidelines for reporting systematic reviews and meta-analyses (Shamseer et al., 2015) and the protocol for this systematic review was registered on INPLASY (https://doi.org/10.37766/inplasy2020.4.0101).

### Review Questions

There are primary, secondary and tertiary questions to be addressed by this review:

#### I. Primary

a. Does prophylactic CQ or HCQ reduce the risk of SARS-CoV-2 infection?
b. Does prophylactic CQ or HCQ reduce the severity of COVID-19 in those subsequently infected, as measured by range or symptoms and laboratory or radiologic abnormalities, need or duration of hospitalization and ICU stay, and subsequent mortality?

#### II. Secondary

a. What are the comparative efficacies of various dosage regimen (choice of CQ versus HCQ, dose strength and duration of treatment) for prophylactic CQ or HCQ in specific at risk populations against SARS-CoV-2 and COVID-19?
b. What is the compliance rate and what are the adverse effects of prophylactic CQ or HCQ at various dose strengths and treatment durations?

#### III. Tertiary

a. What is the quality of the preventative trials assessing CQ and HCQ as prophylaxis against SARS-CoV-2 and COVID-19?
b. How could such trials be optimized to allow better future assessment of the intervention outcomes?
c. How can the results of such trials potentially inform health policy in the pandemic?

### Search method and selection procedure

The main search resources will be 23 national, regional and international clinical trial registries including the WHO International Clinical Trials Registry Platform (Table 1). The search period will be up to the present date of the search. The following search terms will be used: (“covid-19” OR “covid 19” OR “2019-nCoV” OR “n-cov” OR “sars-cov-2” OR “sars-cov2” OR “2019-ncov” OR “SARS-Coronavirus-2” OR “SARS-Coronavirus2”) AND (“chloroquine” OR “hydroxychloroquine”). We will eliminate any duplicates records from different registries and record the reasons for exclusion of trials at various stages of the search as well as outline the selection process in a PRISMA flow diagram. For all included trials, the source clinical trial registry database will be searched and the related trial record identified in order to supplement data extraction. Additional corroborative searches will be executed using Pubmed, Cochrane Central Register of Controlled Trials (CENTRAL), and Embase, and without any language or publication status restrictions. The Infectious Diseases Data Observatory (IDDO) website which documents clinical trial registrations related to COVID-19 (Maguire & Guérin, 2020) will also be consulted. In order to identify articles that might have been missed in the electronic searches, we will a) scan the reference bibliographies of other pertinent systematic reviews on the search terms, and evaluate in full text all the articles they include, b) scan the reference lists of selected narrative reviews and other documents relevant to the subject, c) conduct cross-citation search in Google Scholar, as well as review relevant news websites in English and Chinese for any newly announced or unregistered trials. Grey literature searching will also be conducted for technical or research reports of planned, active or completed clinical trials from industry, international and government agencies, and scientific research groups.

**Table 1.**
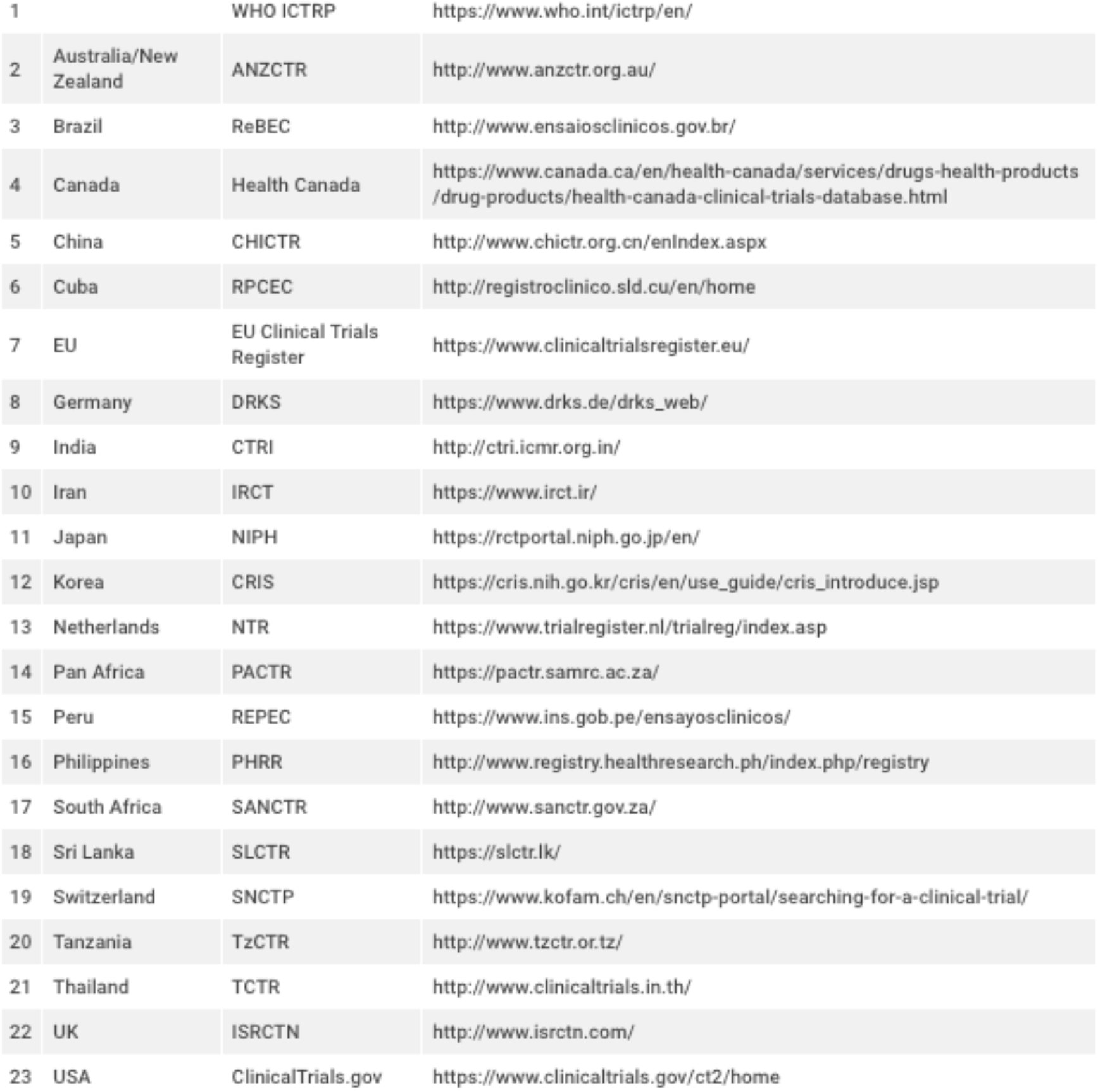
List of source clinical trial registries

### Eligibility criteria

Study eligibility criteria for this systematic review and meta-analysis will be assessed in accordance with established Participants, Interventions, Comparisons, Outcomes and Study designs (PICOS) descriptions (Guyatt et al., 2011):

#### I) Participants

This will include healthy but at-risk subjects who otherwise are without contraindications to participate, as defined by the authors of the trials. Studies including subjects already infected with SARS-CoV-2 will be excluded.

#### II) Interventions

The interventions are the preventative use of CQ or HCQ alone or in combination with other prophylactic agents. We will not restrict our criteria to any dosage, duration, timing or route of administration.

#### III) Comparisons

The comparison will be placebo or other agents chosen by authors (CQ or HCQ with or without other prophylactic agent versus placebo and other same prophylactic agent) or no treatment (CQ or HCQ versus observation). Trials assessing CQ or HCQ plus other agents will be eligible if co-interventions are identical in both intervention and comparison groups.

#### IV) Outcomes

We will not use outcomes as an exclusion criteria during the selection process, but will include all outcomes included by authors of the trials grouped under primary or secondary outcomes. Examples of primary outcomes include seroconversion, incidence and prevalence of subsequent clinical diagnosis and laboratory confirmation of SARS-CoV-2 infection or COVID-19 disease, subsequent severity of COVID-19 disease, hospitalization rate, ICU admission rate, death rate and loss of work-hours (or number of sick days) in those newly infected after intervention is undertaken. Examples of secondary outcomes include side-effects of active intervention and compliancy rates. Such primary and secondary outcomes can be presented in a ‘Summary of Findings’ table, and a table with all the outcomes can be presented in an appendix.

#### V) Study Design

Only randomized controlled trials (RCT) will be included.

### Data extraction

Using standardised forms, two researchers will independently extract data on study design, setting, participant characteristics, intervention and comparison details including dosage, duration, timing and route of administration, outcomes assessed and time of measure, as well as funding source or conflicts of interests as reported by authors of the trials. In the case of the need for further clarification on trial details, we will directly contact the principle investigators of the trials. We will resolve disagreements by discussion, and an independent arbiter will adjudicate any unresolved disagreements.

### Quality and risk of bias assessment

Two researchers will independently assess risk of bias for each RCT study using the Jadad scale (Oxford Quality Scoring System)(Clark et al., 1999). This is composed of five points in total; two for randomization, two for blinding, and one for the drop out rate, and gives an output in reference to the quality of the trial. In case of discrepancy between the two researchers, a third party will be asked to apply the scale to independently address the discrepancy. Additionally, researchers intend to qualitatively summarize the risk of bias across different studies for each of six domains: (1) random sequence generation (2) allocation concealment (3) blinding methods (4) incomplete outcome data (5) selective outcome reporting (6) other biases, as referenced by the Cochrane collaboration network (Higgins et al., 2011).

### Data synthesis and statistical analysis

If there are more than one trial and they are clinically homogeneous, we will conduct meta-analysis using RevMan 5.3 (Nordic Cochrane Centre, 2014), using the inverse variance method with random effects model. For any outcomes where data was insufficient to calculate an effect estimate, a narrative synthesis will be employed. For binary outcomes, we will summarize using risk ratio (RR) and 95% CI. For continuous outcomes, we will use mean difference (MD) and standard deviation (SD) to summarise the data using a 95% CI. The Mantel-Haenszel method (Suesse & Liu, 2019) will be used to pool effect estimates of binary outcomes and inverse variance for continuous outcomes. Cochrane Q test will be used to assess heterogeneity between studies (Higgins et al., 2003), and I^2^ testing will be done to quantify heterogeneity between studies (Higgins & Thompson, 2002), with values > 50% representing moderate-to-high heterogeneity. Either a random or fixed-effect model will be used to pool the data depending on the level of heterogeneity detected and the number of studies involved. Subgroup analysis will also be performed to identify possible causes of significant heterogeneity between studies. In case we identify significant differences between subgroups (test for interaction <0.05), we will report these results separately. If there are at least 10 trials available to be included in this study, we will conduct funnel plot and Egger test to check for reporting bias (Sterne et al., 2011)

## Dissemination of information

The current protocol will be revised periodically and adapted as necessary in accordance with updated RCTs assessing CQ and HCQ prevention in the changing context of the COVID-19 pandemic, and updates will be accessible online via the Inplasy registry. Results of the baseline review as well as updates will be published in as preprint and submitted to an open source, peer-reviewed journal.

## Study status

At the time of protocol submission, preliminary searches and piloting of the study selection process have been completed. A database has been established and data extraction is currently being piloted and tested.

## Data Availability

No data are associated with this article

## Acknowledgements

Not applicable.

## Abbreviations

COVID-19: coronavirus 2019
CQ: chloroquine
HCQ: hydroxychloroquine
RCT: randomized controlled trial
SARS-CoV-2: severe acute respiratory syndrome coronavirus-2

## Authors’ contributions

The study protocol was conceived by RC. RC prepared and drafted the protocol. RC and WZS provided input into the design, edited, revised and approved the protocol.

## Ethics approval and consent for publication

Not applicable

## Registration

This protocol has been registered on INPLASY (INPLASY202040101) and is available at https://doi.org/10.37766/inplasy2020.4.0101.

